# Brain age is longitudinally associated with sensorimotor impairment and mild cognitive impairment in subacute stroke

**DOI:** 10.1101/2024.12.18.24319276

**Authors:** Octavio Marin-Pardo, Mahir H. Khan, Stuti Chakraborty, Michael R. Borich, Mayerly Castillo, James H. Cole, Steven C. Cramer, Miranda R. Donnelly, Emily E. Fokas, Niko H. Fullmer, Jeanette R. Gumarang, Leticia Hayes, Hosung Kim, Amisha Kumar, Emily. A. Marks, Emily R. Rosario, Heidi M. Schambra, Nicolas Schweighofer, Grace C. Song, Myriam Taga, Bethany P. Tavenner, Carolee Winstein, Sook-Lei Liew

## Abstract

**BACKGROUND:** Brain age, a proxy of overall brain health estimated from structural neuroimaging, has been associated with sensorimotor performance in chronic stroke. Similarly, post-stroke cognitive outcomes have been associated with accelerated brain aging. However, the relationships between brain age, sensorimotor, and cognitive outcomes in early subacute stroke (<3 months after onset) are less understood.

**METHODS:** In this work, we investigated associations between stroke survivors’ brain-predicted age difference (brain-PAD, quantified as a person’s brain age minus their chronological age) and longitudinal measurements of motor impairment (Fugl-Meyer Upper Extremity Assessment [FMUE]) and cognitive impairment (Montreal Cognitive Assessment [MoCA]) in subacute stroke. We used high-resolution T1-weighted MRIs from 44 participants at baseline and three months after stroke onset to investigate associations between brain-PAD, MoCA, and FMUE scores with robust linear mixed-effects regression models and mediation analyses.

**RESULTS:** We found negative associations between baseline brain-PAD and FMUE at baseline (β=-0.87, p=0.029) and three months (β=-0.87, p=0.011). Baseline brain-PAD was also negatively correlated with MoCA at three months (β=-0.13, p=0.015) but not at baseline (β=-0.11, p=0.141). Baseline brain-PAD was not associated with changes in FMUE (β=-0.01, p=0.930) or MoCA (β=-0.03, p=0.579). Finally, MoCA was not associated with FMUE at either time point, nor did it mediate the relationship between brain-PAD and FMUE.

**CONCLUSION:** Overall, we show that baseline brain age predicts both motor and cognitive outcomes at three months. However, motor and cognitive outcomes are not directly associated with one other. This suggests that brain age is representative of changes in multiple, distinct neurological pathways post-stroke. Further research with longer time intervals is needed to examine whether brain age also predicts chronic stroke outcomes.

## Introduction

Stroke is a leading cause of long-term adult physical disability and is associated with an increased risk of acquired cognitive decline.^1^ Previous research suggests that accelerated neurodegeneration and brain atrophy post-stroke correlate with motor and cognitive impairments in the chronic phase (>6 months after onset).^2,3^ However, it is unclear how stroke influences the development and evolution of these deficits, particularly in the early months following the stroke.

Brain age, a neuroimaging-based measure, aims to estimate a person’s age using measures of structural brain morphometry and whole brain integrity.^4,5^ As existing brain tissue is crucial for post-stroke neural reorganization and recovery,^6^ brain age has been suggested as a promising biomarker of post-stroke outcomes. Specifically, by calculating the difference between a person’s predicted brain age minus their chronological age (brain predicted age difference, or *brain-PAD*), researchers have found that younger-appearing brains (i.e., lower brain-PADs) are associated with better motor outcomes in chronic stroke.^3,7^ Brain aging has also been linked to numerous neurodegenerative processes^8^ and has been shown to be accelerated after stroke and in those with Alzheimer’s disease,^9,10^ with younger-appearing brains having a lower risk of developing cognitive impairment up to three years after stroke^11^ and better motor outcomes.^3,7^

Although researchers have shown that concurrent motor and cognitive impairments can be present after a mild stroke,^12^ it is unclear how these impairments may be related to each other and to brain age in early subacute stroke (<3 months after onset). Therefore, the goal of this work was to investigate associations between stroke survivors’ brain-PAD and measurements of motor and cognitive impairment in the subacute phase of recovery. We hypothesized that older-appearing brains (higher brain-PAD) would be associated with both more severe motor (lower FMUE) and cognitive impairments (lower MoCA). Furthermore, we hypothesized that such deficits would be associated via both direct and indirect (mediated) relationships.

## Methods

The primary and corresponding authors of this study had full access to the data and take responsibility for data integrity and analyses. As detailed below, we used a publicly available brain age model,^13^ which can be found at https://photon-ai.com/enigma_brainage. Code used to extract and format FreeSurfer features of interest can be found at https://github.com/npnl/ENIGMA-Wrapper-Scripts. Additional data and code from this study are available upon reasonable request from the corresponding author.

We conducted a prospective, longitudinal study across three sites, collecting 1-mm isotropic T1-weighted brain structural magnetic resonance images (MRI), demographic data, and clinical assessments for sensorimotor impairment (Fugl-Meyer Upper Extremity assessment [FMUE]) and mild cognitive impairment (Montreal Cognitive Assessment [MoCA]). We recruited adult stroke survivors in the subacute phase of recovery and acquired brain imaging and behavioral data at baseline and around three months after stroke onset. Participants eligible for this study presented with arm weakness in at least one major muscle group (e.g., with a score of less than 5 points in the Manual Muscle Test). Participants were excluded if they had a prior history of traumatic brain injury, a major musculoskeletal or secondary neurological condition, or a MoCA score below 10 points (indicating severe cognitive impairment).^14^ Written informed consent was obtained from all participants in accordance with the Declaration of Helsinki. The University of Southern California Health Sciences Campus Institutional Review Board and the local ethics board of each cohort (Casa Colina Hospital, Emory University, and NYU Langone Health) approved to conduct this study.

Data curation and processing were conducted following established pipelines and protocols.^3,15^ Briefly, MRI data was processed using the FreeSurfer longitudinal reconstruction pipeline (recon-all, v. 7.4.1) to automatically segment 153 neuroanatomical features (e.g., bilateral cortical thicknesses, cortical surface areas, subcortical volumes), which then were used to calculate individual brain ages using a validated and publicly available machine-learning model.^13^ Stroke lesions were manually segmented by trained researchers, normalized, and registered to the MNI-152 template.^16^ Then, lesion volume metrics were extracted using the Pipeline for Analyzing Lesions after Stroke toolbox.^17^ Finally, we investigated cross-sectional and longitudinal associations between brain-PAD, FMUE, and MoCA scores in separate robust linear mixed-effects regression models including intracranial volume, lesion volume, sex, and age as fixed effects and cohort as random effect (Models 1 to 5; where Δ represents the difference between values at three months minus their corresponding baseline values). Additionally, we performed a 3-step mediation analysis^18^ by examining 1) whether brain-PAD was associated with FMUE, 2) whether brain-PAD was associated with MoCA, and 3) whether MoCA had an indirect, mediating effect on the relationship between baseline brain-PAD and FMUE. All statistical analyses were performed using R (v. 4.4.1). The following libraries were used to organize, test, and analyze the data: *tidyverse*, *reshape*, *lme4*, *car*, *influence.ME*, *diptest*, *robustlmm*, and *mediation*.

FMUE ∼ BrainPAD_baseline_ + ICV + Lesion Volume + Sex + Age + (1|Cohort) Model 1

ΔFMUE ∼ BrainPAD_baseline_ + ICV + Lesion Volume + Sex + Age + (1|Cohort) Model 2

MoCA ∼ BrainPAD_baseline_ + ICV + Lesion Volume + Sex + Age + (1|Cohort) Model 3

ΔMoCA ∼ BrainPAD_baseline_ + ICV + Lesion Volume + Sex + Age + (1|Cohort) Model 4

FMUE ∼ MoCA_baseline_ + ICV + Lesion Volume + Sex + Age + (1|Cohort) Model 5

## Results

We examined 44 participants from three cohorts. Table 1 presents a summary of participant demographics. We performed linear mixed-effects regressions and evaluated residuals plots, quantile-quantile plots, variance inflation factors, and the presence of influential observations in the data. Because influential outliers were found in the regression models examined, as determined using Cook’s distance, we used robust linear mixed-effects regression models to maximize the dataset by reducing the weight of influential observations without removing them.^19^ We also removed influential outliers (identified as noted above) to perform the mediation analysis using linear mixed-effects regressions. As customary in brain age analyses, we confirmed the association of predicted brain ages and chronological ages and found positive correlations between them at baseline (r=0.69, p<0.001) and three months (r=0.65, p<0.001). Changes in brain-PAD, FMUE, and MoCA were statistically significant for the group, with participants showing increased brain aging and improved FMUE and MoCA scores over time, as shown in Table 1 and Figure 1.

**Figure 1.**
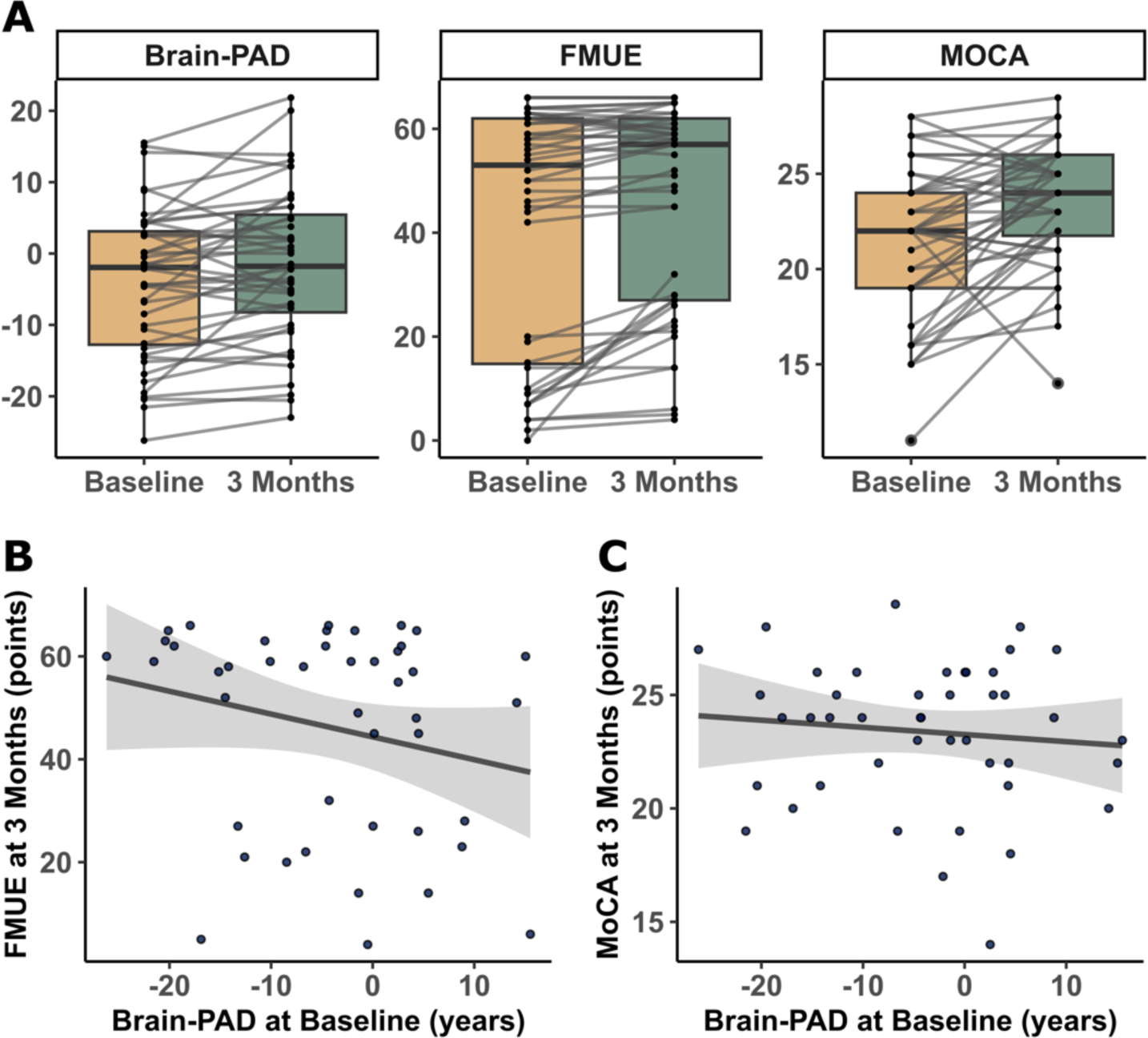
Data Distributions **(A)** Brain predicted age difference (brain-PAD; t=3.04, p=0.004; baseline mean=-4.07±10.40, 3-month mean=-1.67±10.72), Fugl-Meyer Assessment of the Upper Extremity (FMUE; t=3.90, p<0.001; baseline mean=41.98±23.72, 3-month mean=46.16±19.95), and Montreal Cognitive Assessment (MoCA; t=3.87, p<0.001; baseline mean=21.41±4.09, 3-month mean=23.39±3.19) distributions at baseline and three months after onset. Boxplots show group medians in black and individual trajectories of each participant in gray. **(B)** Trend lines for the association between baseline brain-PAD and FMUE at 3 months (β=-0.87, p=0.011). **(C)** Trend lines for the association between baseline brain-PAD and MoCA at 3 months (β=-0.13, p=0.015).

**Table 1.**
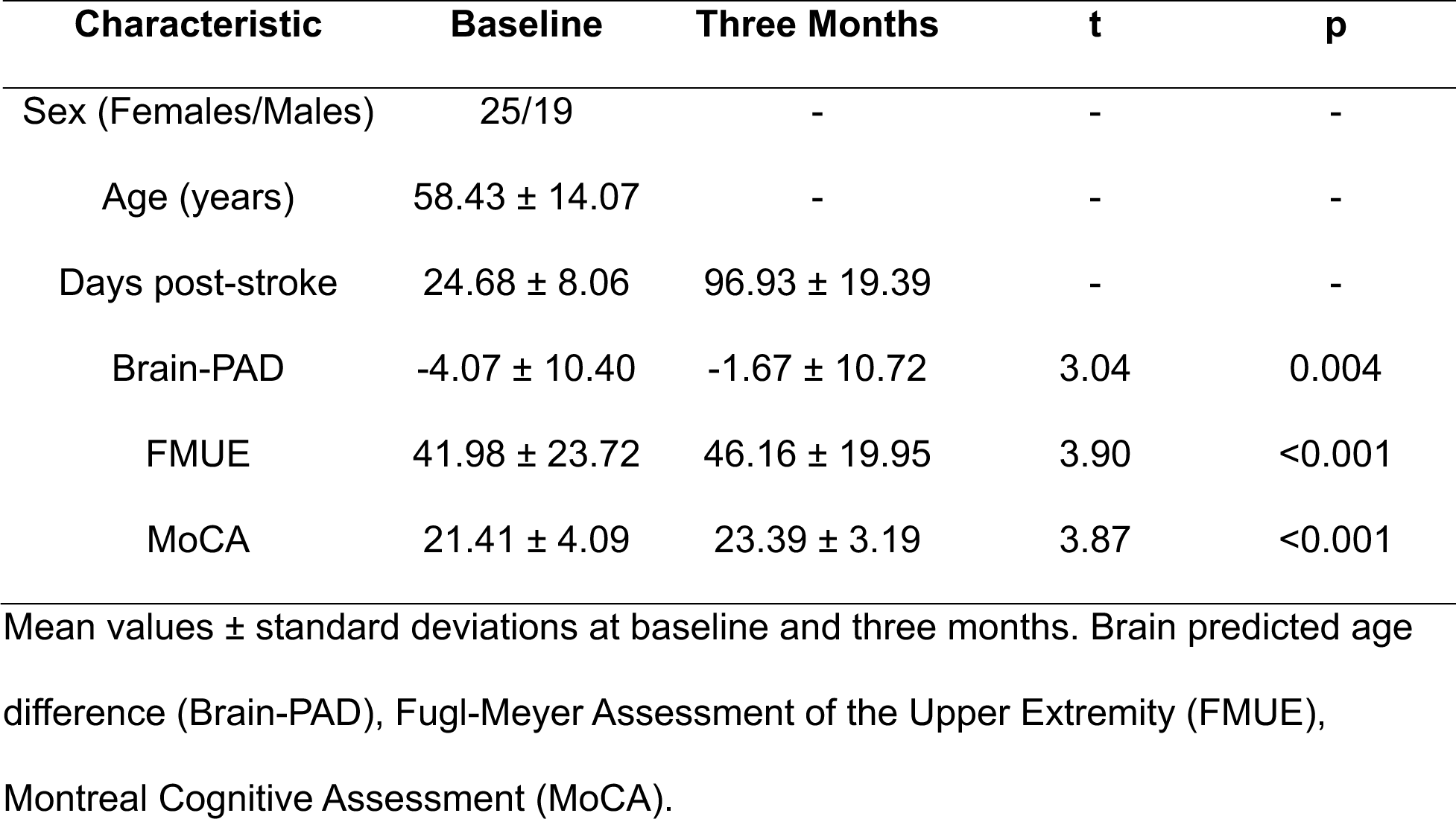
Participant demographics and changes in clinical assessments and Brain-PAD.

| Characteristic | Baseline | Three Months | t | p |
| --- | --- | --- | --- | --- |
| Sex (Females/Males) | 25/19 | - | - | - |
| Age (years) | 58.43 ± 14.07 | - | - | - |
| Days post-stroke | 24.68 ± 8.06 | 96.93 ± 19.39 | - | - |
| Brain-PAD | -4.07 ± 10.40 | -1.67 ± 10.72 | 3.04 | 0.004 |
| FMUE | 41.98 ± 23.72 | 46.16 ± 19.95 | 3.90 | <0.001 |
| MoCA | 21.41 ± 4.09 | 23.39 ± 3.19 | 3.87 | <0.001 |

Mean values ± standard deviations at baseline and three months. Brain predicted age difference (Brain-PAD), Fugl-Meyer Assessment of the Upper Extremity (FMUE), Montreal Cognitive Assessment (MoCA).

We found negative associations between baseline brain-PAD with FMUE scores at baseline (β=-0.87, p=0.029, 95% CI −1.64 to −0.09) and at three months (β=-0.87, p=0.011, 95% CI −1.54 to −0.20) (Model 1), as shown in Figure 1 and Table 2. However, baseline brain-PAD was not associated with changes in FMUE (β=-0.01, p=0.930, 95% CI −0.18 to 0.16) (Model 2).

**Table 2.** Relationships between sensorimotor and cognitive outcomes and Brain-PAD.

| *Predictors* | FMUE (Three Months), $R^2=0.68$ | | | | MoCA (Three Months), $R^2=0.33$ | | | |
| --- | --- | --- | --- | --- | --- | --- | --- | --- |
| | $\beta$ | SE | 95% CI | *p* value | $\beta$ | SE | 95% CI | *p* value |
| **Brain-PAD** | **-0.87** | **0.34** | **-1.54 – -0.20** | **0.011** | **-0.13** | **0.05** | **-0.24 – -0.03** | **0.015** |
| ICV | -0.32 | 3.82 | -7.81 – 7.17 | 0.933 | 0.35 | 0.61 | -0.85 – 1.55 | 0.568 |
| Lesion Vol. | -4.79 | 4.32 | -13.26 – 3.69 | 0.268 | -0.17 | 0.67 | -1.49 – 1.14 | 0.795 |
| Sex | -2.10 | 7.36 | -16.53 – 12.34 | 0.776 | 0.03 | 1.19 | -2.30 – 2.37 | 0.978 |
| **Age** | -0.37 | 0.26 | -0.88 – 0.14 | 0.154 | **-0.13** | **0.04** | **-0.21 – -0.05** | **0.002** |

Summary statistics from robust mixed-effects linear regressions to test associations
between baseline brain-PAD and FMUE at three months and baseline brain-PAD and MoCA at three months. Conditional $R^2$ , $\beta$ , SE, 95% CI, and *p* value for all fixed-effect covariates are reported. Significant predictors are denoted in bold. Brain-predicted age difference (Brain-PAD), Fugl-Meyer Assessment of the Upper Extremity (FMUE), intracranial volume (ICV), lesion volume (Lesion Vol.), Montreal Cognitive Assessment (MoCA).

Baseline MoCA was not associated with brain-PAD (β=-0.11, p=0.141, 95% CI −0.25 to 0.04) at baseline but was associated with age (β=-0.13, p=0.022, 95% CI −0.24 to −0.02) (Model 3). Baseline brain-PAD was negatively associated with MoCA scores (β=-0.13, p=0.015, 95% CI −0.24 to −0.03) and age (β=-0.13, p=0.002, 95% CI −0.21 to −0.05) at three months (Model 3). However, baseline brain-PAD was not associated with changes in MoCA (β=-0.03, p=0.579, 95% CI −0.15 to 0.08) (Model 4).

Baseline MoCA was not associated with FMUE at baseline (β=-0.34, p=0.715, 95% CI - 2.17 to 1.49) nor at three months (β=-0.12, p=0.881, 95% CI −1.74 to 1.60) (Model 5). Furthermore, we did not find a mediation effect of MoCA on the relationship between baseline brain-PAD and FMUE at baseline (β=0.05, p=0.587) or three months (β=0.06, p=0.482).

## Discussion

We aimed to investigate the relationships between brain aging, motor outcomes, and cognitive outcomes in the subacute phase of stroke recovery. Our results suggest that older-appearing brains at baseline predict worse sensorimotor and cognitive outcomes three months after stroke onset. This is in line with previous research showing that stroke survivors with older-appearing brains may be less resilient to stroke damage, resulting in long-term sensorimotor impairment,^3^ and have a higher risk of developing cognitive impairment in the chronic phase.^11^ However, further research is necessary to better understand the role of the stroke lesion extent and location on regional changes in brain aging, particularly in subacute stroke.

In addition to investigating cross-sectional and longitudinal associations between brain aging and behavioral outcomes, we also sought to investigate whether sensorimotor and cognitive deficits were associated with one another. Our results suggest that cognitive impairment is not directly associated with motor impairment at baseline or three months after stroke. Previous research has shown that, although both impairments can occur simultaneously after a stroke,^12,20^ the role of cognitive function on motor performance and recovery is less understood.^21^ Previous work has shown that cognitive impairment in the acute phase predicts disability, as measured by the Rankin scale, in the subacute phase after mild stroke.^20^ Similarly, researchers have found that different domains of cognitive function are associated with motor performance after mild subacute stroke.^12^ Since cognitive impairment after stroke has been shown to depend on additional factors (e.g., demographics, comorbidities, lesion characteristics),^1,22^ future work could further study the influence of these and other covariates on the relationship between motor and cognitive outcomes.

Finally, although we hypothesized that cognitive impairment may mediate the relationship between brain aging and sensorimotor impairment, we did not find this to be true. Instead, brain-PAD, which may be a more global construct reflecting the whole brain’s health, is associated with both. Together, our results suggest that the overall integrity of the brain, which is crucial for neural plasticity mechanisms^6^ and overcoming known consequences of stroke (e.g., diaschisis),^23^ is critical in generally supporting recovery across distinct domains.

One possible limitation of this study is the uneven distribution of the FMUE scores we observed, which was bimodal in nature and could limit the generalizability of our findings. Although this was mitigated by using robust regression, future research using a larger, more diverse sample could help to better understand the influence of this and additional factors (e.g., clinical history, lesion characteristics, additional neuroimaging-based markers) in brain aging and behavioral outcomes.

Overall, this research suggests that brain aging might be a promising biomarker of subacute motor and cognitive impairment after stroke. These insights may contribute to widening our understanding of stroke recovery and aid in developing precision rehabilitation tools to improve diagnosis and treatment.

## Data Availability

As detailed in the manuscript, we used a publicly available brain age model, which can be found at https://photon-ai.com/enigma_brainage. Code used to extract and format FreeSurfer features of interest can be found at https://github.com/npnl/ENIGMA-Wrapper-Scripts. Additional data and code from this study are available upon reasonable request from the corresponding author.

